# Bayesian approach for modelling the dynamic of COVID-19 outbreak on the Diamond Princess Cruise Ship

**DOI:** 10.1101/2020.06.21.20136465

**Authors:** Chao-Chih Lai, Chen-Yang Hsu, Hsiao-Hsuan Jen, Amy Ming-Fang Yen, Chang-Chuan Chan, Hsiu-Hsi Chen

## Abstract

The outbreaks of acute respiratory infectious disease with high attack rates on cruise ships were rarely studied. The outbreak of COVID-19 on the Diamond Princess Cruise Ship provides an unprecedented opportunity to estimate its original transmissibility with basic reproductive number (R_0_) and the effectiveness of containment measures. The traditional deterministic approach for estimating R_0_ is based on the outbreak of a large population size rather than that a small cohort of cruise ship. The parameters are therefore fraught with uncertainty. To tackle this problem, we developed a Bayesian Susceptible-Exposed-Infected-Recovery (SEIR) model with ordinary differential equation (ODE) to estimate three parameters, including transmission coefficients, the latent period, and the recovery rate given the uncertainty implicated the outbreak of COVID-19 on cruise ship with modest population size. Based on the estimated results on these three parameters before the introduction of partial containment measures, the natural epidemic curve after intervention was predicted and compared with the observed curve in order to assess the efficacy of containment measures. With the application of the Bayesian model to the empirical data on COVID-19 outbreak on the Diamond Princess Cruise Ship, the R_0_ was estimated as high as 5.71(95% credible interval: 4.08-7.55) because of its aerosols and fomite transmission mode. The simulated trajectory shows the entire epidemic period without containment measurements was approximately 47 days and reached the peak one month later after the index case. The partial containment measure reduced 34% (95% credible interval: 31-36%) infected passengers. Such a discovery provides an insight into timely evacuation and early isolation and quarantine with decontamination for containing other cruise ships and warship outbreaks.

## 1 Introduction

The ongoing pandemic of coronavirus disease 2019 (COVID-19), caused by severe acute respiratory syndrome coronavirus 2 (SARS-CoV-2), was first identified in Wuhan, Hubei, China in December 2019 (WHO, 2020; Huang et al., 2020; Guan et al., 2020; Li et al., 2020), and then declared as a Public Health Emergency of International Concern (PHEIC) on Jan. 30^th^, 2020 and a global pandemic on Mar. 11^st^, 2020 (WHO, 2020). To contain this rapid and wide spread of COVID-19 outbreak, it is imperative to quantify correctly the basic reproductive number (R_0_) defined as the average number of secondary cases generated from a primary case, a useful indicator of infectious disease transmission, locally and globally. We can use R_0_ larger or smaller than 1, being determined by the contact rate, transmission probability, and infectious period, to assess the possibility of potential spread or the control of COVID-19 in affected communities (Ferguson et al., 2005).

At the beginning of COVID-19 epidemic, several studies have tried to quantify R_0_ by the models based on parameters derived from the SARS epidemic in 2003 (Wu et al., 2020; Imai et al., 2020a; Imai et al., 2020b), which was estimated as 2-3, a value close to the R_0_ of SARS (Raily et al., 2003). As the epidemic evolves over time, the difference between SARS and COVID-19 has been gradually noted in terms of transmission mode and infectious period in relation to clinical symptoms and signs (Huang et al., 2020; Guan et al., 2020; Li et al., 2020; Wang et al., 2020). The property of asymptomatic infection and various clinical symptoms and signs of COVID-19 render the force of spread intractable to be estimated by the SARS models. The disparity of transmission between SARS and COVID-19 has been further speculated by the rampant spread in the Washington and the New York states of USA with a doubled increase from Mar. 26^th^ to Apr. 1^st^ and also in the beginning epidemics of European countries such as Italy and Spain (WHO, 2020).

The COVID-19 outbreak on the Diamond Princess Cruise Ship (NIID, 2020a; NIID, 2020b) provides an unprecedented opportunity for us to understand its original transmissibility and force of spread by three unique epidemic-related characteristics. First, susceptible, contact, transmission, and infectious period are clearly defined according to a fixed cohort of global passengers and crews living in the isolated setting of this cruise ship. Second, such a transmission mode through the isolated environment of the cruise ship provides strong human-to-human community-acquired infection in close setting distinct from the transmission mode through household or hospital and relevant healthcare institutions while assessing the transmissibility of this emerging infectious disease. The third is that the infected persons onboard consist of multi-nationals giving us a better sense of the transmissibility of COVID-19 across racial groups. Based on these three characteristics related to the transmission mode on the cruise ship, two thorny questions that have been not addressed in the previous studies are raised (Mizumoto et al., 2020; Sawano et al., 2020; Rocklöv et al., 2020). Can we estimate and predict the epidemic curve to determine timely containment measures and evacuation? What is the effectiveness of containment measures on quarantine and isolation for the infected passengers before evacuation? Answering these two questions plays a crucial role in the guidance of how and when containment measures can be designed and offered for the coming cruise ships that are susceptible to the outbreak of COVID-19.

Although the conventional deterministic approach by fitting the compartment models with the ODE system to observed number of reported cases can shed light on the dynamic of COVID-19 transmission and R_0_ at population level, such an approach may not feasible for the outbreak occurring on cruise ship with modest size of susceptibles. To cope with the uncertainty of outbreak in a confined environment, we thus developed a Bayesian compartment models for the COVID-19 outbreak on the Diamond Princess Cruise Ship.

The rest of the article is organized as follows. Section 2 briefs the empirical data on the outbreak of COVID-19 on the Diamond Princess Cruise Ship alone with the events took place in the quarantine measures on board. Section 3.1 gives the details on the specification of Bayesian ODE compartment model depicting the propagation of COVID-19 through the states of susceptibles, exposed, infectives, and removed. The methods for validating the fitted cases and evaluating the efficacy of containment measures are addressed in Section 3.2. Section 4 shows the estimated results on the dynamic of COVID-19 outbreak on the Diamond Princess Cruise Ship by fitting the Bayesian ODE compartment model to the empirical data. Section 5 gives the discussion of applications and implications on the proposed Bayesian ODE compartment model to assess the disease outbreak data.

## 2 Empirical data

### 2.1 Data on COVID-19 outbreak of the Diamond Princess cruise ship

The data on the voyage of Diamond Princess cruise ship, the reported number of confirmed COVID-19 cases, and the quarantine process were retrieved from publicity available sources including the Princess Cruise website of the Carnival Cooperation (Princess Cruise Lines, 2020), the official website of Ministry of Health, Labor and Welfare, Japan (Ministry of Health, Labor and Welfare, Japan, 2020), and the National Institute of Infectious Disease, Japan (NIID, 2020a; NIID, 2020b). The auxiliary information sources including the general media and scientific news and web-based summaries were also used for a cross-check on the cases numbers and the quarantine measures and the evacuation after the outbreak in Diamond Princess cruise ship (Princess Cruise Lines, 2020; Wikipedia, 2020a; Wikipedia, 2020b; Garcia-Navarro, 2020; Normile, 2020). A prospective observational study design on the propagation of the COVID-19 outbreak was applied to evaluating the transmissibility of SARS-CoV-2. Details on the derivation of information on COVID-19 outbreak on the Diamond Princess Cruise Ship are provided in the Supplementary material A.

### 2.2 COVID-19 outbreak on the Diamond Princess Cruise Ship

The Diamond Princess Cruise Ship departed from Yokohama on Jan. 20^th^, 2020. There was a total of 3711 subjects including 2666 passengers from 11 countries (1285 from Japan, 470 from Hong Kong, 425 from USA, 215 from Canada, 40 from United Kingdom, 25 from Russia, 20 from Taiwan, 15 from Israel, 13 from New Zealand, and the rest of 158 from Australia and Germany) and 1045 crew members on the voyage of the Diamond Princess (NIID, 2020a; Ministry of Health, Labor and Welfare, Japan, 2020; Wikipedia, 2020a). The outbreak started from a passenger from Hong Kong who had joined with part of the voyage and disembarked on Jan. 25^th^, 2020 who was later confirmed as a COVID-19 case through laboratory testing (Ministry of Health, Labor and Welfare, Japan, 2020; Wikipedia, 2020a). The Diamond Princess Cruise Ship was therefore quarantined at the Yokohama harbor since its dock on Feb. 5^th^, 2020. Following the index case, 10 cases of COVID-19 were reported on Feb. 5^th^, 2020, which heralded the outbreak yielding 762 cases in the following weeks (WHO, 2020; Ministry of Health, Labor and Welfare, Japan, 2020; Wikipedia, 2020a). Up to Feb. 29^th^, the COVID-19 outbreak on the Diamond Princess Cruis Ship have resulted in 6 deaths (WHO, 2020).

### 2.3 Measures of containment for COVID-19 transmission on the Diamond Princess Cruise Ship

During the quarantine process, the personal protective equipement was provided for crew members. Passangers and crews were instructed to monitor body temperature by themselves. Subjecs with body temperature higher than 37.5 degree celsus or developed any illness were referred to the medical center on the ship where medical services were provided and the test for SARS-CoV-2 were referred. To maintain operations of the ship, some crew continued to perform essential but limited services while the ship remained in quarantine (NIID, 2020a). This symptom-based approach started since the ship dock at Yokohama harbored on Feb. 5^th^. With the gradual expansion of laboratory capacity from Feb. 11^st^, the quarantine officers provided test systematically gauged by testing capacity and covered passengers without the manifestation of symptoms from Feb. 14^th^ (NIID, 2020a; NIID, 2020b).

**Figure 1** shows both the epidemic curve and cumulative cases of all passengers on the Diamond Princess Cruise Ship during Jan. 20^th^ to Feb. 29^th^, 2020. The cases were identified mainly based on the symptom of fever and the close contacts defined by sharing the cabin with confirmed cases during the quarantine process followed by laboratory test (NIID, 2020a; NIID, 2020b; Ministry of Health, Labor and Welfare, Japan, 2020). Passengers who were tested positive for SARS-CoV-2 were removed from the ship and transferred to health care facilities on land. Subjects with negative test result will be kept in the ship for the rest of quarantine period. The COVID-19 cases identified on the Diamond Princess Cruise Ship were reported on daily basis by the Ministry of Health, Labor and Welfare, Japan (2020). On Feb. 16^th^, 2020, several countries started requesting the evacuation of their citizens from the cruise ship. The USA was the first country to pull out 380 Americans on Feb. 17^th^, 2020. It should be noted that the quarantine measure has commenced from Feb. 5^th^ but isolation and decontamination were gradually expanded from Feb 14^th^ until the complete evacuation on Feb. 19^th^, 2020 (NIID, 2020a; NIID, 2020b).

**Figure 1.**
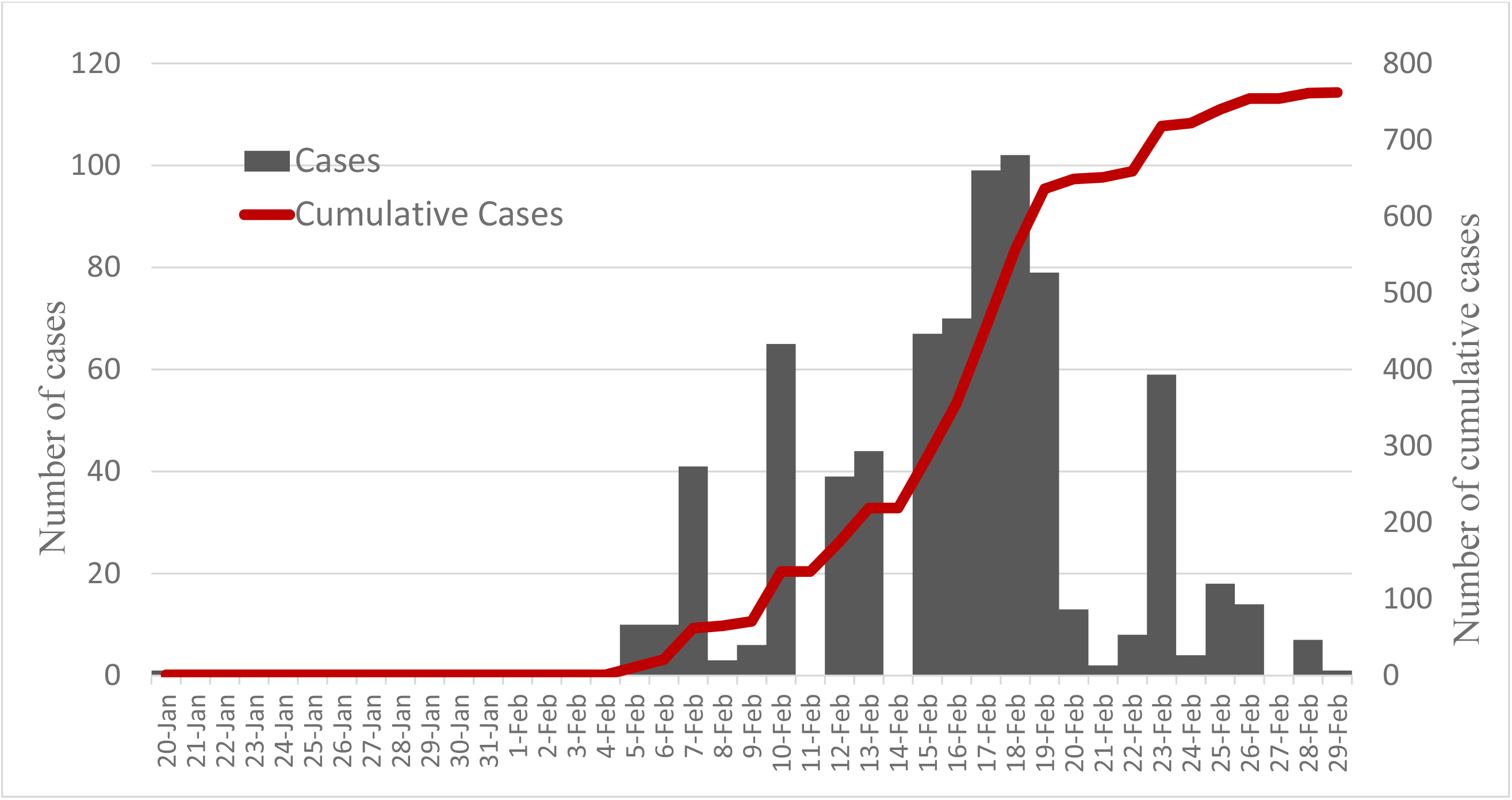
The epidemic curve and cumulative cases of COVID-19 on the Diamond Princess Cruise Ship. The daily reported COVID-19 cases (gray bar, left Y-axis) and cumulative cases (red line, right Y-axis) on the Diamond Princess Cruise Ship from Jan. 20th to Feb. 29th, 2020.

The data used for the following analysis consisted of the cases reported between Jan. 20^th^ to Feb. 19^th^, 2020, the date of ending quarantine announced by Ministry of Health, Labor and Welfare, Japan. Detailed counts of the cases and the information sources are provided in Supplementary Material **S-Table 1**. The positive rate for SARS-CoV-2 among passengers tested was 20.6% (621/3011). Among 621 SARS-CoV-2 infected cases, 322 were asymptomatic (51.9%, **S-Table 1** in the Supplementary Material).

## 3 Bayesian compartment model for the dynamic of COVID-19 outbreak

### 3.1 Model specification

To predict the propagation of COVID-19 outbreak on the cruise ship making use of the observed number of cases with the consideration of uncertainty, a Bayesian ODE SEIR model was developed. A four-compartment model (**Figure 2**) was applied to describe the daily flow of the population on the Diamond Princess cruise ship between the four status with respect to COVID-19 infection, including S (susceptible population), E (exposed persons to infectious persons), I (infectious persons with COVID-19), and R (recovered persons from infectious status or infectious persons being isolated or removed from the cruise ship, or dead) states.

**Figure 2.**
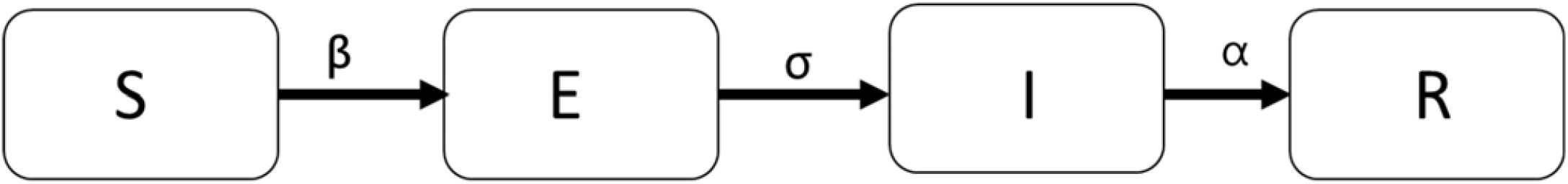
The SEIR (Susceptible, Exposure, Infective, and Recovered) model for the evolution of COVID-19 in Diamond Princess Cruise Ship.

Let *s*(*t*), *e*(*t*), *i*(*t*), and *r*(*t*) denote the numbers of susceptibles, exposed, infectives, and removed/isolated/recovered/dead on the cruise ship at time *t*. The four-compartment system can be depicted by using the following ordinary differential equations to describe the instantaneous change of the four types with time

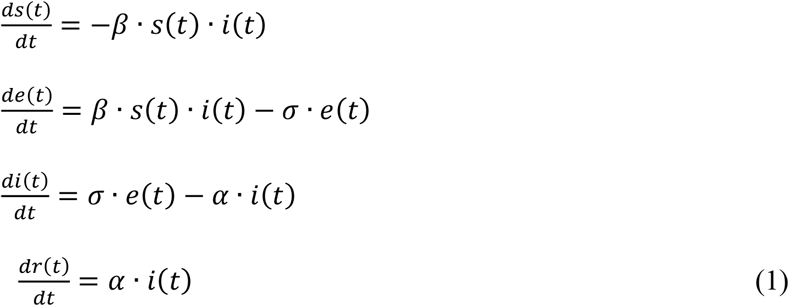

With the solution of the four-state differential equations regarding the empirical data on the observed COVID-19 cases in Diamond Princess cruise ship, the parameters of transmission coefficient (*β*), progression rate (*σ*) to and recovery rate (*α*) from the infectious status can be estimated. The estimated results were further used to depict the propagation on the number of four COVID-19 infective states by time. Note that because the population of the Diamond cruise ship is fixed, the following equation always holds.

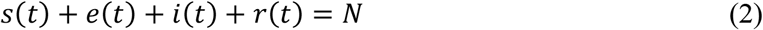

where *N* is the initial size of this cohort on lockdown, namely the 3711 subjects being quarantined in Diamond Princess cruise ship.

The Bayesian Markov Chain Monte Carlo method for the ordinary differential equation system for the four-compartment SEIR model was used to estimate parameters of interests. The Bayesian approach facilitates us to incorporate the prior information of parameters. As COVID-19 is an emerging disease, little is known about the time interval between exposed and infected, as well as between infected and recovery. We borrowed the experience from the first report in Wu-Han that is the origin infectious resource for the outbreak of the Diamond Princess cruise ship. We then assigned Gamma(53, 278) and Gamma(24, 168) for σ and α in order to fit the mean duration from exposed to infective and from infective to recovery for 5 (95% CI: 4-7) and 7 (95% CI: 5-12) days, respectively (Huang et al., 2020; Guan et al., 2020).

Since only data on the number of reported confirmed cases per day were available, they contain those already infected, regardless of appearance of clinical symptoms and signs, and recovered cases, we therefore modeled two random variables, number of unknown infected status and the confirmed cases per day following two normal distributions of

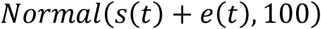

and

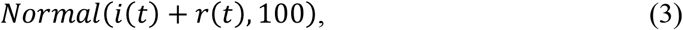

respectively. Following the derivation of transmission coefficient, *β*, the basic reproduction number (R_0_) were estimated with function of transmission coefficient (*β*) and recovery rate (*α*) as follows,

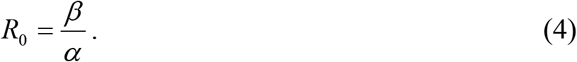

The Bayesian Markov Chain Monte Carlo method for the ordinary differential equation system for the four-compartment SEIR model was used to estimate parameters of interests. A series of Bayesian ODE SEIR models were applied to quantify the force of COVID-19 spreading by using the data up to Feb. 19^th^, 2020.

### 3.2 Model validation and prediction of COVID-19 dynamic on the Diamond Princess Cruise Ship

To assess the homogeneity of estimated results on transmission coefficient and basic reproductive number derived from Bayesian ODE compartment model across the quarantine period, data up to Feb. 13^th^ (before systemic test), Feb. 16^th^ (before the evacuation of passengers from USA), and Feb. 19^th^ were used. The validation of the model was assessed by comparing the fitted cumulative COVID-19 cases with the observed number. The effectiveness on the quarantine measures with the consideration of evacuation was further assessed based on the estimated result derived from the period between Jan. 10^th^ to Feb. 19^th^ in conjunction with the extended data after the evacuation to predict the cases up to Feb. 29th, 2020. The evaluation of efficacy was derived by the comparison with the total observed cases up to Feb. 29th excluding the index case from Hong Kong.

## 4 Results

### 4.1 Transmissibility of SARS-CoV-2 on cruise ship

By using a Bayesian ODE SEIR model in conjunction with reported cases series, we estimated the transmission coefficient and R_0_ for the COVID-19 outbreak on the Diamond Princess Cruise Ship in **Table 1**. The R_0_ of was estimated as 5.71 (95% credible interval (CI): 4.08 − 7.55) based on the cases up to Feb. 19^th^, 2020. Consistent results were derived when different time points for the observed epidemic with the estimated R_0_ ranging between 5.65 to 5.67 with overlapping 95% CIs. The SEIR model demonstrated a satisfactory fit by using the information on data prior to the Feb-19, 2020 as shown in **Figure 3 and Table 2**.

**Table 1.** Estimated basic reproductive numbers of SARS-CoV-2. The results on the transmission coefficient (β) and R0 estimated from the Bayesian ODE SEIR model based on three epochs between Jan. 20th to Feb. 19th. The estimated results on the transmission coefficient (β) shows consistent result of 0.21. Three corresponding basic reproductive number estimates were 5.67 (95% CI: 3.93-7.98), 5.65 (95% CI: 3.95-7.42), and 5.71 (95% CI: 4.08-7.55) of three ending epochs.

| Scenario | Parameter | Estimate | 95% Credible interval |
| --- | --- | --- | --- |
| **Up to 2020-02-13** | Basic reproductive number ($R_0$) | 5.67 | (3.93, 7.68) |
| | Transmission coefficient ($\beta$) | 0.21* | (0.16, 0.27) |
| **Up to 2020-02-16** | Basic reproductive number ($R_0$) | 5.65 | (3.95, 7.42) |
| | Transmission coefficient ($\beta$) | 0.21* | (0.16, 0.26) |
| **Up to 2020-02-19** | Basic reproductive number ($R_0$) | 5.71 | (4.08, 7.55) |
| | Transmission coefficient ($\beta$) | 0.21* | (0.17, 0.26) |

\* $\times 10^{-3}$

**Table 2.**
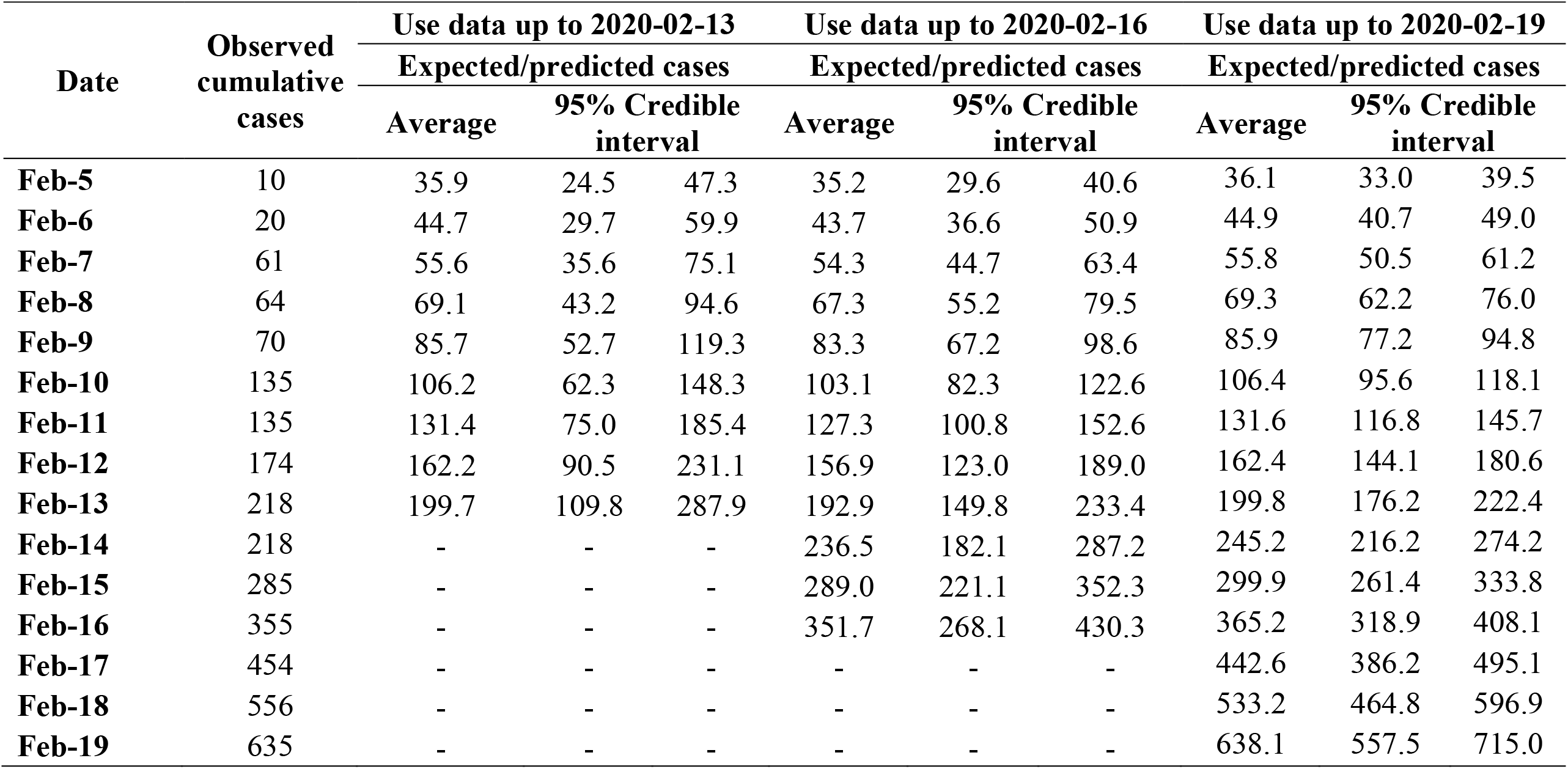
Observed and expected cumulative COVID-19 cases by date using data up to designated dates.

| Date | Observed<br>cumulative<br>cases | Use data up to 2020-02-13 |  |  | Use data up to 2020-02-16 |  |  | Use data up to 2020-02-19 |  |  |
| --- | --- | --- | --- | --- | --- | --- | --- | --- | --- | --- |
|  |  | Expected/predicted cases |  |  | Expected/predicted cases |  |  | Expected/predicted cases |  |  |
|  |  | Average | 95% Credible<br>interval |  | Average | 95% Credible<br>interval |  | Average | 95% Credible<br>interval |  |
| **Feb-5** | 10 | 35.9 | 24.5 | 47.3 | 35.2 | 29.6 | 40.6 | 36.1 | 33.0 | 39.5 |
| **Feb-6** | 20 | 44.7 | 29.7 | 59.9 | 43.7 | 36.6 | 50.9 | 44.9 | 40.7 | 49.0 |
| **Feb-7** | 61 | 55.6 | 35.6 | 75.1 | 54.3 | 44.7 | 63.4 | 55.8 | 50.5 | 61.2 |
| **Feb-8** | 64 | 69.1 | 43.2 | 94.6 | 67.3 | 55.2 | 79.5 | 69.3 | 62.2 | 76.0 |
| **Feb-9** | 70 | 85.7 | 52.7 | 119.3 | 83.3 | 67.2 | 98.6 | 85.9 | 77.2 | 94.8 |
| **Feb-10** | 135 | 106.2 | 62.3 | 148.3 | 103.1 | 82.3 | 122.6 | 106.4 | 95.6 | 118.1 |
| **Feb-11** | 135 | 131.4 | 75.0 | 185.4 | 127.3 | 100.8 | 152.6 | 131.6 | 116.8 | 145.7 |
| **Feb-12** | 174 | 162.2 | 90.5 | 231.1 | 156.9 | 123.0 | 189.0 | 162.4 | 144.1 | 180.6 |
| **Feb-13** | 218 | 199.7 | 109.8 | 287.9 | 192.9 | 149.8 | 233.4 | 199.8 | 176.2 | 222.4 |
| **Feb-14** | 218 | - | - | - | 236.5 | 182.1 | 287.2 | 245.2 | 216.2 | 274.2 |
| **Feb-15** | 285 | - | - | - | 289.0 | 221.1 | 352.3 | 299.9 | 261.4 | 333.8 |
| **Feb-16** | 355 | - | - | - | 351.7 | 268.1 | 430.3 | 365.2 | 318.9 | 408.1 |
| **Feb-17** | 454 | - | - | - | - | - | - | 442.6 | 386.2 | 495.1 |
| **Feb-18** | 556 | - | - | - | - | - | - | 533.2 | 464.8 | 596.9 |
| **Feb-19** | 635 | - | - | - | - | - | - | 638.1 | 557.5 | 715.0 |

**Figure 3.**
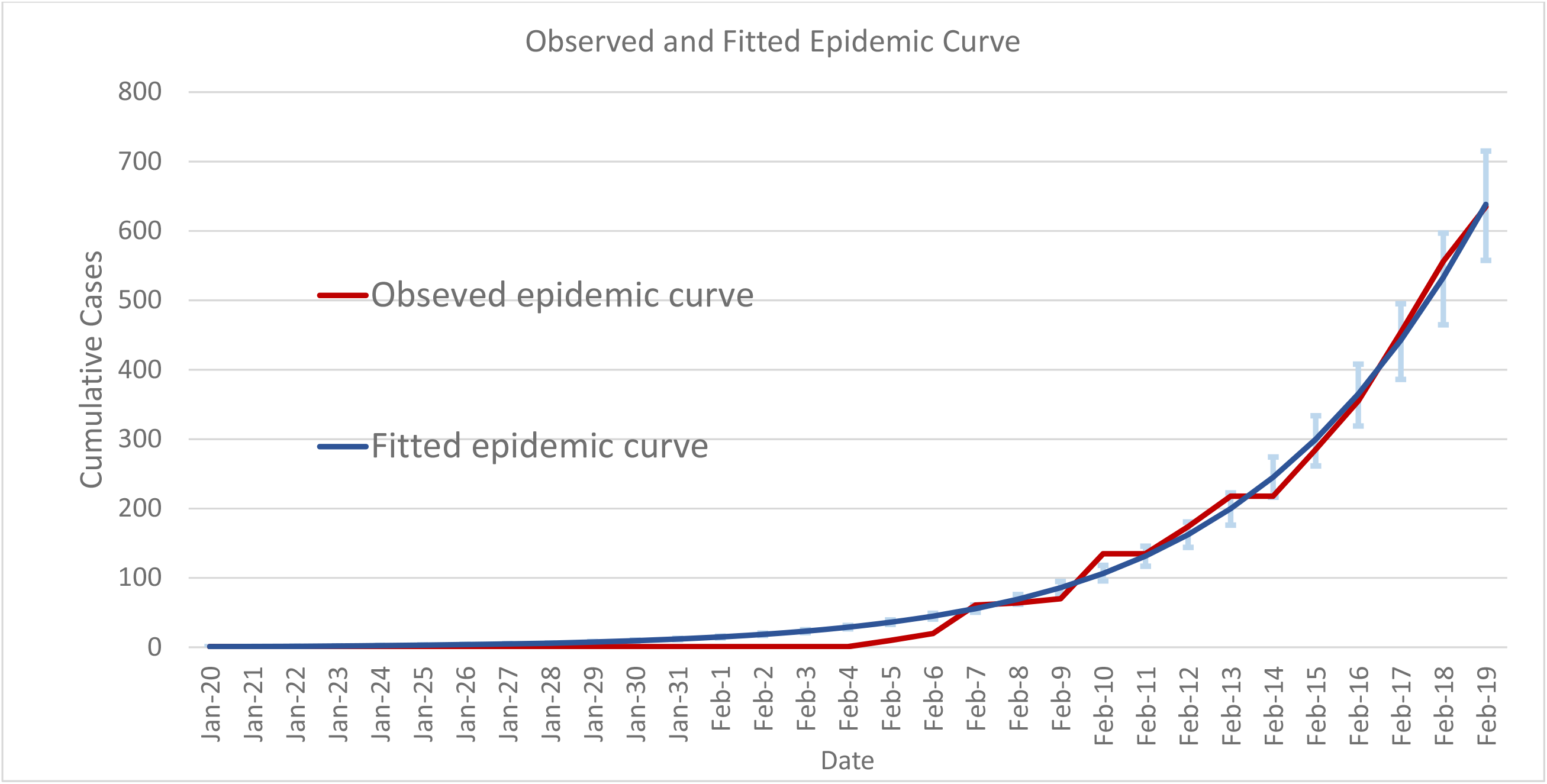
The fitted cases versus the observed cases for the COVID-19 outbreak on the Diamond Princess Cruise Ship based on the Bayesian ODE SEIR model. The observed curve (red line) is close to the SEIR-fitted epidemic curve (blue line with error bar standing for 95% credible interval on the frequency of fitted cases each day).

**Figure 3** shows the observed cases as opposed to the fitted ones on cumulative cases derived from the Bayesian ODE SEIR model with the detailed fitted cumulated cases listed in **Table** 2. Based on the transmission parameter estimated by the empirical data from Jan. 20^th^ to Feb. 19^th^, the fitted cumulated cases on Feb. 19^th^ are 638 (95% CI: 558-715), which is comparable to the observed cumulative cases of 635 (χ^2^_(1)_ = 0.02, *P*=0.90).

### 4.2 The dynamic of COVID-19 on cruise ship

Based on the parameters of basic reproductive number trained by the data before Feb. 19^th^, **Figure 4(a)** shows the predicted cumulative curve given the scenario of no containment measures to control the COVID-19 outbreak on the Diamond Princess Cruise Ship with the detailed daily predicted counts listed in **S-Table 2** in the Supplementary Material. While the total of 3135 (95% CI: 3001-3258) were infected and only 16% passengers remain susceptible the outbreak would be expected to end on Mar. 7^th^ according to the theory of herd immunity when more than 82% [(1-1/5.71)×100%] of total population have been infected. The evolution of COVID-19 outbreak can further be decomposed into the status of susceptible, exposed, infected, and removed (including recovery and death) among the 3711 passengers of the Diamond Princess Cruise Ship. **Figure 4(b)** shows this dynamic epidemic curve in terms of these four components. The passengers were exposed to an increasing risk of being infected by SARS-CoV-2 and further developed into COVID-19 during the period of Jan. 20^th^ to Feb. 26^th^ which reached the peak of 976 (95% CI: 770-1200) on Feb. 26^th^. On Feb. 19^th^, 2020, the number of subjects with effective exposure and infected with SARS-CoV-2 without the manifestation of clinical symptom for COVID-19 was estimated as 603 (95% credible interval (CI) 443-775) (**S-Table 3** in the Supplementary Material), while the COVID-19 cases were predicted as 638 (95% CI: 558-715) (**S-Table 2** in the Supplementary Material) consisting of 369 (95% CI: 298-453) infected cases and 270 (95% CI: 200-344) removed cases (**S-Table 3** in the Supplementary Material). Based on this dynamic epidemic curve, 50% of passengers were infected by SARS-CoV-2 until Feb. 22^nd^ (**S-Table 3** in the Supplementary Material). During the acceleration period of the outbreak (from Jan 10^th^ to Feb. 26^th^), the exposed subjects (**Figure 4(b)**, yellow line) were more than the infected (**Figure 4(b)**, blue line). The difference between two components was estimated as 234 on Feb. 19^th^ (**S-Table 3** in the Supplementary Material). The corresponding figures were 121 (52% of Feb. 19^th^) and 70 (30% of Feb 19^th^) on Feb 14^th^ and 11^st^, respectively (**S-Table 3** in the Supplementary Material). This difference represents the expected number of subjects who have been exposed but not confirmed as infected cases yet would mix up with the susceptibles during evacuation. This accounted for the additional 126 confirmed cases requiring quarantine and isolation after the evacuation of those already exposed and still susceptible. The dynamic curve suggests Feb. 26^th^, the crossed point between the exposed and the infected curve as shown in **Figure 4(b)**, may be a better evacuation time of the remaining uninfected passengers and crews than the real evacuation date of Feb. 19^th^.

**Figure 4.**
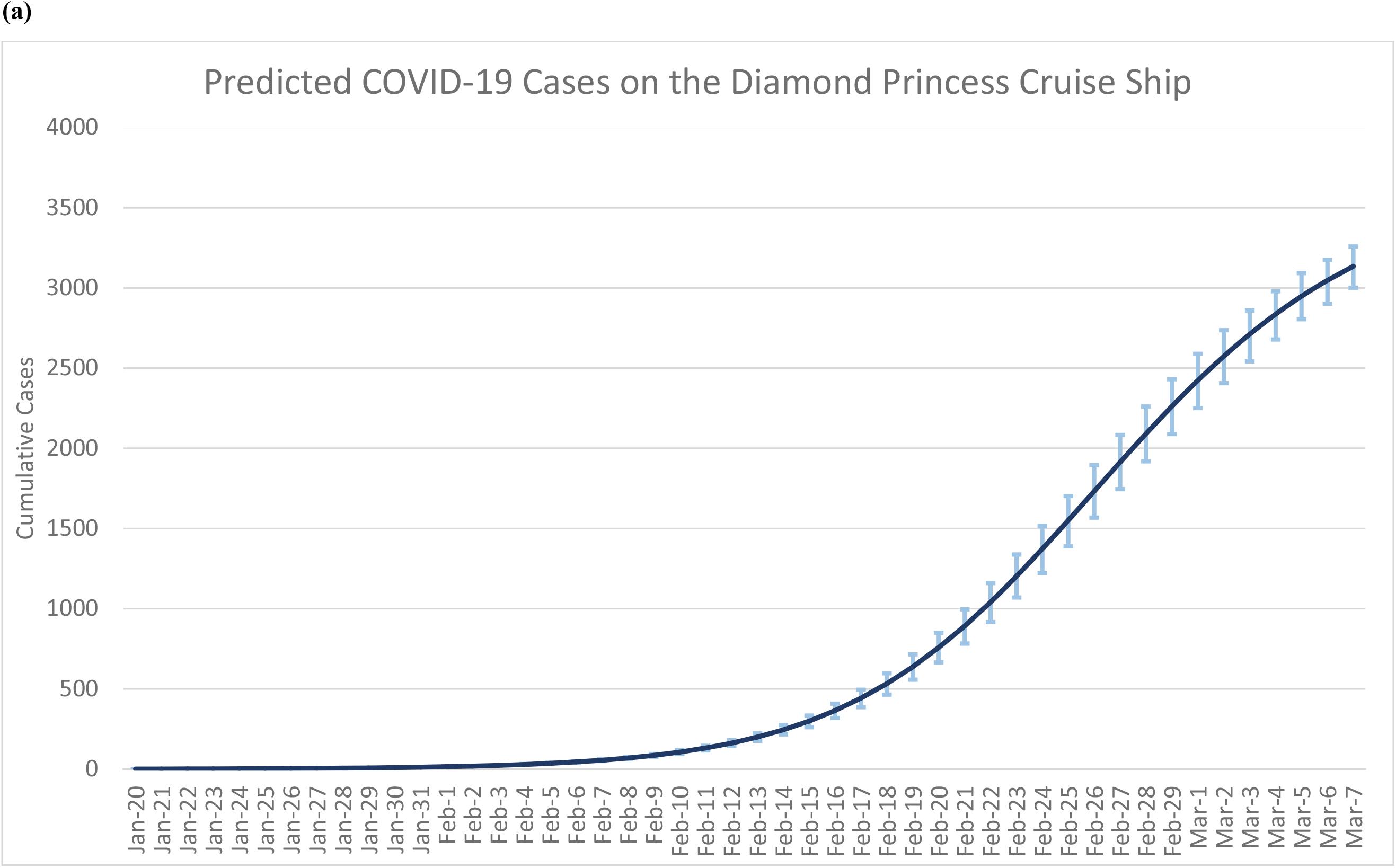
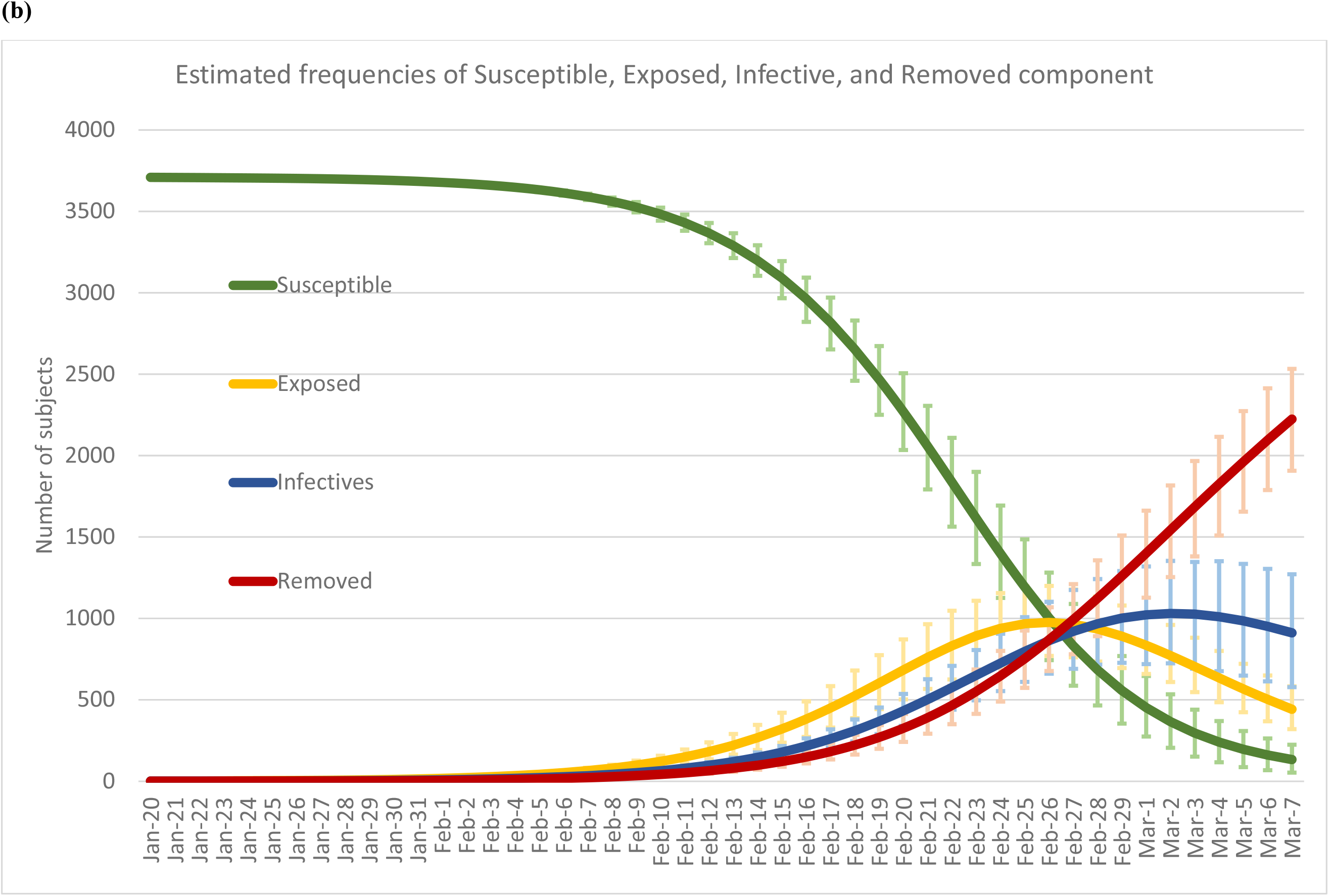
Predicted COVID-19 cases. Figure 4(a) shows the predicted number of daily cumulated COVID-19 cases that occurred on the Diamond Princess Cruise Ship given the scenario of no containment measures till Mar. 7^th^. Figure 3(b) shows the dynamic curves of the SEIR model, including the frequencies of the components of susceptible (S, green line), exposed (E, yellow line), infectious (I, blue line), and removed (R, red line) along with the estimated 95% credible intervals on the dynamics of each component.

### 4.3 Efficacy of containment measures

Based on the estimated results on the transmissibility of SARS-CoV-2 derived from the observed data on COVID-19 outbreak, **Figure 4** shows the effectiveness of containment measures implemented on the Diamond Princess Cruise Ship. The total number of COVID-19 cases associated with the outbreak on the Diamond Princess Cruise Ship on Feb. 29^th^ was 761 (**Figure 5**, red line). Compared with the scenario of no containment measures (2262.4, 95% CI: 2088.4-2431.1, **Figure 5**, blue line) the effectiveness of quarantine and partial isolation in reducing the infected passengers was 34% (95% CI: 31-36%) based on the estimate of relative risk of 0.66 (95% CI: 0.64-0.69) between the observed and the expected cases.

**Figure 5.**
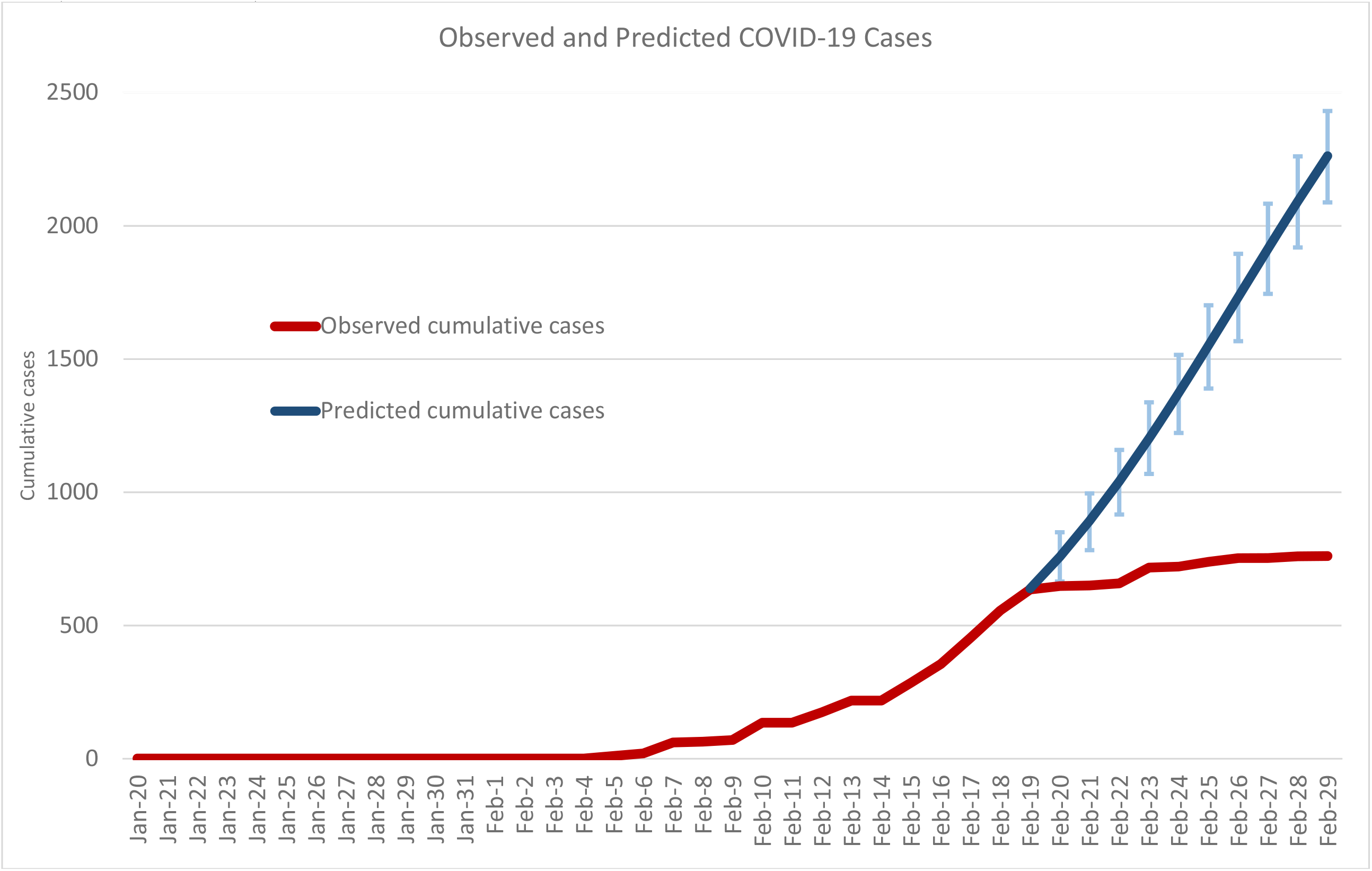
Effectiveness of containment measures on quarantine and isolation. The blue line shows the expected cumulative cases without containment measure on the Diamond Princess Cruise Ship. The red line is the observed daily cumulative case with containment measure. The total number of COVID-19 cases associated with the outbreak on the Diamond Princess Cruise Ship at Feb. 29^th^ was 761. Compared with the scenario of no intervention (2262.4, 95% CI: 2088.4-2431.1) the effectiveness of quarantine and isolation in reducing the infected passenger was 34% based on the estimate of relative risk 0.66 (95% CI: 0.64-0.69) between the observed and the expected.

## 5 Discussion

Due to a lack of preparedness on quarantine and isolation that have been often seen in the beginning stage of early epidemic on a new pathogen, the outbreak of COVID-19 in such a confined situation with the characteristics of fixed cohort and multi-national passengers provides a natural opportunity to estimate the transmissibility of this new SARS-CoV-2 and also insight into the deployment of containment measures for the outbreak of other cruise ships that are susceptible to COVID-19. It is very striking to see that the basic reproductive number in such a scenario was estimated up to 5.71 (95% CI: 4.08-7.55), which is unusually high compared with the transmissibility of SARS with the estimated R_0_ ranging from 2 to 3 (Riley et al., 2003), given its aerosols and fomite transmission mode because the persistence of SARS-CoV-2 on inanimate surfaces were reported.^22^ More importantly, this unusually high value of R_0_ is reproducible in the beginning of several epidemics of COVID-19 such as Korea, Iran, and Italy and the subsequent initial epidemics waves in the Europe and in the USA. The high value of R_0_ also accounts for the soaring number of confirmed cases reported in a short period due to the accelerated transmission amplified by a series of clustered events due to public gathering. The recent outbreaks in the Washington and the New York state of USA triggered by a series of clustered events including long-term care facilities follow such a similar pattern (WHO, 2020). With such a high transmissibility, the virus can lead to an overwhelming number of COVID-19 cases and further bring extraordinary stress to health care system as well as social and economic infrastructure.

Based on the simulated projection of natural dynamic epidemic curve without containment measure, the entire infectious process took 47 days from the introduction of index cases since Jan. 20^th^ until the end of epidemics on Mar. 7^th^ when 84% of passengers would be expected to be infected (3135 cases) to reach the phenomenon of herd immunity. Excluding two weeks of the incubation period from the introduction of index case, the epidemic cycle of COVID-19 on the Diamond Princess Cruise Ship is around one month. The natural dynamic curve also provides the control group for the evaluation of the effectiveness of containment measures on quarantine and isolation measures. The partial containment measure on quarantine and isolation in the current scenario of the Diamond Princess led to the reduction of infected passengers by only 34% (95% CI: 31-36%). Had the complete containment measures been implemented on Feb. 14^th^, the total number of cases would have been reduced from 761 to 410 cases, resulting in the effectiveness of 54% (95% CI: 46-64%, **S-Table 4** in the Supplementary Material) and an even larger effectiveness would have been achieved given an early implementation on Feb. 10^th^ suggested from the estimated dynamic curve (**Figure 4(b)**) while the testing capacity is limited. Moreover, even when the implementation was delayed to Feb. 14^th^, the measure adopted to guarantee complete containment as evacuation with adequate deposition should be a high priority. However, timely evacuation is also important as those who were arranged to be evacuated also bore a high risk due to the mixing up of subjects already exposed not yet confirmed as infected cases. The timely evacuation would be expected after the peak of the transmission dynamic on Feb. 26^th^ rather than the real date of Feb. 19^th^. The decision on this early evacuation thus results in the subsequent occurrence of cases among these exposed subjects after evacuation returning to each country that may invoke cluster infection of public gathering and the epidemic in the community. These findings derived from the Diamond Princess can be applied to military ships. COVID-19 cases have been reported on four USA aircraft carriers since the end of March. In the outbreak occurring on the USS Theodore Roosevelt, 1156 COVID-19 cases out of 5000 onboard Navy sailors were reported (Wikipedia, 2020c). Given the similar situation of clustering and close contact in a confined setting, the timely containment measures and evacuation should be implemented as suggested by the transmission dynamic derived from the Diamond Princess outbreak.

The proposed Bayesian ODE SEIR model provides a flexible approach to elucidate the transmission of SARS-CoV-2 by using aggregate data on observed cases with limited resolution. Thanks to the Bayesian framework, the information on disease characteristics such as incubation and transmission period learned from previous studies (Huang et al., 2020; Guan et al., 2020) can be incorporated into our models with their uncertainty being considered. The parameters estimated from the integration of observed data and state-of-the-art knowledge on disease pattern can shed light on the transformation of disease pattern which is crucial for the allocation of resources and the implementation of strategies for us to contain the coming cruise ships that may be attacked by COVID-19.

## Data Availability

Source data are available in the supplementary materials. The code for analysis is available from author upon request.

## Acknowledgments

We thank Professor Stephen Duffy from Center of Cancer Prevention, Wolfson Institute of Preventive Medicine, Queen Mary University of London, Dr. Peter Dean from University of Turku, Finland, and Dr. Smith Robert from American Cancer Society for the discussion and suggestions. This work was financially supported by the “Innovation and Policy Center for Population Health and Sustainable Environment (Population Health Research Center, PHRC), College of Public Health, National Taiwan University” from The Featured Areas Research Center Program within the framework of the Higher Education Sprout Project by the Ministry of Education (MOE) in Taiwan.

## Author contributions

Conceptualization: CCC, and HHC; study design: CCL, CCC, and HHC; methodology: CCL, CYH, and MFY; data retrieval and management: HHJ and MFY; statistical analysis: CYH and MFY; computer programming: HHJ, CYH, and MFY, writing: CCL and CYH, revision of draft: CCC and HHC; All authors agreed the findings and provided input on the revision of the manuscript.

## Competing interests

Authors declare no competing interests

## Funding Support

Ministry of Science and Technology, Taiwan (MOST 108-2118-M-002-002 −MY3; MOST 108-2811-M-002 −640; MOST 108-2118-M-038-001-MY3), Ministry of Education, Taiwan (NTU-107L9003).

## Role of Funder/Sponsor

The funders had no role in the design and conduct of the study; collection, management, analysis, and interpretation of the data; preparation, review, or approval of the manuscript; and decision to submit the manuscript for publication.

## Notes

### Competing Interest Statement

The authors have declared no competing interest.

